# Cytological Abnormalities and its relation to CD4 count among HIV seropositive women living in Ahvaz, southwest of Iran

**DOI:** 10.1101/19004416

**Authors:** Amene Darvishi, Seyed Mohammad Alavi, Morteza Abdullatif Khafaie, Alireza Sokooti, Shahla Molavi, Shokralleh Salmanzadeh

## Abstract

**Introduction:** Human immunodeficiency virus (HIV) infection is a known risk factor for abnormal cervical cytology and cervical cancer. The aim of this study was to investigate cervical cytological abnormalities and its relation with CD4 (T4 Lymphocyte) count among HIV seropositive women.

**Methods:** We conducted a study on 58 HIV positive women referred to Ahvaz Counseling Center for Behavioral Disease, southwest of Iran between 2016 and 2017. Pap smear was performed for all participants from the cervix and endocervix. Patient’s’ characteristics including age, duration of disease, treatment with anti-retroviral treatment (ART), marital status, number of children, and contraception method were also recorded. Cervical cytological abnormalities reported as Bethesda system (TBS). A regular blood sample was taken from all the patients to evaluate the CD4 cells counts. Logistic regression models were used to obtain OR of presences of cytological abnormalities related to CD4 counts, controlling for important factors.

**Results:** Out of 58 patients only 5 were not under ART. We demonstrated that 29.3 % of patients had squamous cell abnormalities and these abnormalities, was more prevalent among 30-40 years old patients (70.6%). The prevalence of ASC-US (Atypical Squamous Cells of Undetermined Significance), LSIL (Low-Grade Squamous Intraepithelial Lesions) and HSIL (High-Grade Squamous Intraepithelial Lesions) were 19.0%, 3.4%, and 6.9% respectively. Overall 9 patients need to repeat Pap smear test. Presence of cervical cytological abnormalities was not associated with the CD4 count, even after adjusting for the variable such age, duration of disease and ART.

**Conclusion:** We found a high prevalence of ASC-US in HIV-infected women which was independent of age, duration of diseases and history of ART. Though cervical cancer screening in this population might have a substantial public health benefit.

**Summary box:**

- More than 70% of cervical cancers incidences associated with Genital HPV infections
- Prevalent of Squamous cell abnormalities among HIV-infected women was about sex time more than general population
- We demonstrated that squamous cell abnormalities are more prevalent in middle age women (30 to 40 years)
- The high prevalence of Squamous cell abnormalities in HIV-infected women warrants the need for regular Pap smear screening

## Introduction

Acquired immune deficiency syndrome (AIDS) still is one of the major causes of death worldwide[1, 2] and cancer is the leading cause of death in these population[3]. Since the primary demonstration of Harold zur Hausen in the early 1980s, the link between genital HPV infections and cervical cancer are well established[4, 5] and accountable for 70% to 80% of advanced cervical cancers[6, 7].

About half a million new cervical cancer and 266,000 death occurred in 2012 worldwide[8]. According to the WHO and the International Agency for Research on Cancer (IARC), cervical cancer is the second most common cancer in women in low and middle-income countries[9, 10].

Partially the differences in the occurrence of disease in developed and developing nation could be due to the absence of screening and lifestyle changes programs, insufficient infrastructure and trained personnel[10, 11]. Women with HIV are at greater risk of developing squamous cell adhesion (SIL) and cervical intraepithelial neoplasia (CIN) and cervical cancer[12]. Several studies have shown that the incidence of CIN and cervical cancer in women with HIV is about 2 to 22 times higher than that of HIV-negative women[13, 14]. Cervical epithelial cell abnormalities (ECA) represent a range of SIL from mild to severe dysplasia to aggressive concessions[15]. Unfortunately, despite the existing recommendations, most of the HIV-positive women are not screened properly[16] and a large number of deaths are due to delayed detection of cervical cancer[12].

Cervical carcinoma is a preventable disease through early cytological and cytotoxic screening that indicates the role of Pap smear[13, 17]. Women in advanced stages of HIV (Stage III and IV), having multiple sexual partners and long-term contraceptive pills (OCP), have increased the risk of ECA[13, 18]. Immunological status such low CD4 levels also play an important role in cervical carcinogenesis[13].

The present study was conducted to evaluate the cytological changes in the pap smear and its association with the CD4 count. We hypothesis that prevalence of abnormal Pap smear among HIV seropositive women is higher than healthy population and CD4 count linked with this situation.

## Methods

This cross-sectional study was done among a cohort of fifty eight HIV seropositive women above 18 years old. Subjects attending Ahvaz Counseling Center for Behavioral Diseases between 20^th^ March 2016 and 19^th^ March 2017 recruited for the study after the Ethical Committee’s approval. All patients were informed and written consent was taken before enrolment. The diagnosis of HIV was based on the presence of two HIV Ab ELISA positive tests and confirmation by western blot test. Pregnant women, subject with the irregular periodic cycle, virgin, and those with a history of cervical dysplasia and cervical cancer were excluded from the study (see figure 1).

**Figure 1:**
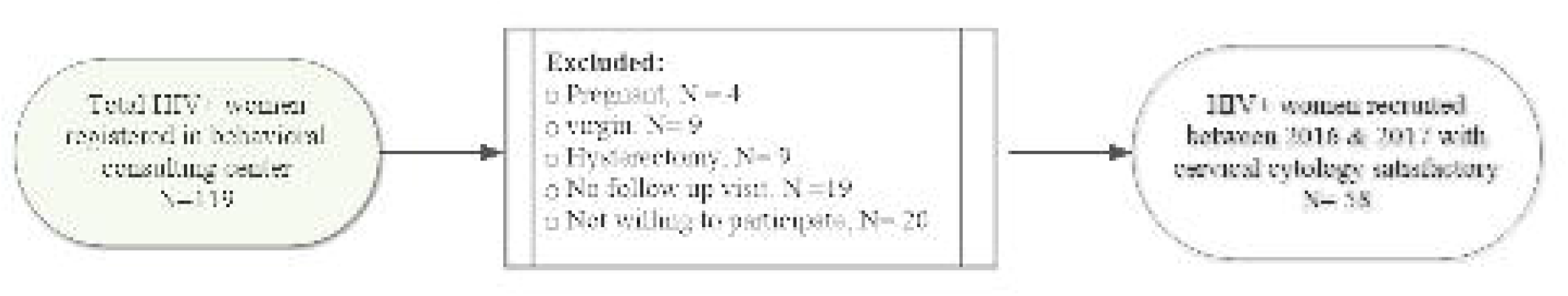
The flowchart displays the selection of the overall HIV seropositive women enrolled in the study. exclusions are shown

### Patient and Public Involvement

We did not involve patients or the public in our work. Socio-demographic information including age, duration of disease, marital status, number of children, mode of HIV transmission, contraception method, and history of ART (at least a month treatment assumed to be under treatment) was collected. A blood sample was taken from all the patients on completion of the morning visit. CD4+ lymphocytes count determined using CyFlow miniPOC which is reliable and accurate[19] and reported as cells in a microliter (cells/μl).

### Cytology sampling

Pap smear tests conducted by a trained midwife and all procedure performance were under the responsibility of a gynecologist from Department of Obstetrics and Gynecologist, Razi hospital, Ahvaz, Iran. The exocervix and endocervix samples were taken with Cervex-Brush^®^ Combi in accordance with the standard protocol[20]. Brush disconnected from the handles, and the heads were deposited into a BD Sure Path™ ethanol-based preservative fluid, which can be stored for weeks at room temperature. Samples vial capped, labeled and delivered to Pasteur lab on the liquid-based media. In the laboratory, blood cells and inflammatory exudates were removed from the suspension and the samples homogenized. The smears were confectioned on slides and stained using BD PrepStain slide processor. All samples were interpreted and reported by a pathologist according to the Bethesda system 2001[21] which is comparable with 2014 with a minor revision[22]. In the current study the cervical cytological interpretation was denominated as negative for intraepithelial lesions and malignancy (NILM) which includes inflammatory changes, organisms, atrophic changes and reactive changes; atypical squamous cells of undetermined significance (ASC-US), cannot exclude HSIL (ASC-H), squamous intraepithelial lesions (SILs) including low-grade SILs (LSILs) and high-grade SILs (HSILs) and invasive carcinoma. The presence and severity of inflammation in the smears reported as mild (< 30 inflammatory cells high power field), moderate (30 to 100 inflammatory cells/high power field) and severe (> 100 inflammatory cells/high power field). Prevalence of bacterial vaginosis (BV), Trichomonas vaginitis (TV), and vaginal candidiasis (VC) was determined in the samples. Women with Pap smear abnormality were referred to the Department of Obstetrics & Gynecology as per American Society for Colposcopy and Cervical Pathology[23].

### Statistical Analysis

Characteristics of patients including mean, frequencies, and contingency tables described. Visual inspection of data and Shapiro-Wilk W test were used to investigate normality of variables. We calculated Square-root of CD4 count since it was not normally distributed (i.e. heteroscedastic standard division). Logistic regression was used to find the association between CD4 count and presences of squamous cells abnormality (ASC-US and SIL) and NILM, adjusted for important risk factors. The results are given as absolute change in the CD4 count for presences of cytological abnormality (i.e. ASUS/SIL and NILM). For model selection we investigated possible predictors of CD4 such age, duration of disease, history of ART (yes vs. no) and contraception as an indicator variable for oral contraceptive pill, tubal ligation, and condom) and marital status (married vs. unmarried), using t-test and Pearson’s correlation coefficient, as univariate analysis. The significance threshold level of P=0.05 was used in all analysis. All analysis was carried out using STATA version 13 software (STATA Corporation, College Station, TX).

## Results

A total of 58 patients with satisfactory cervical cytology were included in the study. The patients mean age was 35.26 ranging from 22 to 59 years. Mean duration of HIV infection was 3.57 ± 2/34 years (ranging from 1 to 10 years) and 91.4% of the patients were treated with ART) see table 1).

**Table1:** Characteristics of participated HIV-seropositive women referred to Ahvaz Counseling Center for Behavioral Diseases between 20^th^ March 2016 and 19^th^ March 2017

|  |  |
| --- | --- |
| Age- mean± SD years | 35.26± 7.68 |
| Being under ART- F (%) | 53 (86) |
| Duration of disease- mean± SD years | 3.57± 2.35 |
| Married- F (%) | 54 (93) |
| Parenting at least a child- F (%) | 41 (71) |
| Modes of transmission (i.e. HIV) |  |
| Sexual- F (%) | 46 (79) |
| Contraception, |  |
| OCP/TL- F (%) | 10 (12) |
| Condom- F (%) | 36 (62) |
| CD4 count- median (25<sup>th</sup>- 75<sup>th</sup> percentile) μL | 565 (390- 760) |
| ART= anti retro virus therapy; OCP= Oral contraceptive pill; TL= Tubal Ligation; CD4= CD4-positive T-lymphocytes |  |

### Interpretation of Pap smear

From total satisfactory for evaluation Pap smear 27 case (46.6.4%) endocervical/transformation zone component (EC/TZ) were present (table 2). Forty-one case (71%) were Negative for intraepithelial lesion or malignancy (NILM). Out of NILM, 21 cases (36%) showed normal cytological finding (figure 2a) and, 37 cases (64%) were inflammatory. Two cases had features of fungal organism morphology consistent with Candida Spp (figure 2b). Non-neoplastic cases with metaplasia and atrophic cervicitis were 9 and 2 cases respectively. Normal results without any cytological anomalies, organism and inflammation were observed in 25.9% of patients. Seventeen (29.3%) patients had abnormalities in cervical squamous cells, including 2 (11.7%) low LSILs (figure 1c), 4 (23.5%) High HSILs (figure 2d) and 11 (64.7%) ASC-US (figure 2e). In the present study, most of the anomalies (70.6%) were observed in the age group of 30 to 40 years’ old.

**Table 2:** Categorization of Cytodiagnosis of HIV-seropositive women referred to Ahvaz Counseling Center for Behavioral Diseases

| **Cytodiagnosis** | **No. of patients** | **Percentage** |
| --- | --- | --- |
| Inadequate, EC/TZ | 31 | 53.40 |
| NILM | 41 | 70.70 |
| Normal | 15 | 25.90 |
| Inflammatory | 37 | 63.80 |
| Non-specific | 35 | 60.34 |
| Candida spp. | 2 | 3.45 |
| Non-neoplastic | 11 | 18.96 |
| Metaplasia | 9 | 15.52 |
| Atrophic | 2 | 3.45 |
| ASCUS | 11 | 18.96 |
| SIL | 6 | 10.34 |
| HSIL | 4 | 6.90 |
| LSIL | 2 | 3.45 |
| Need repeat test | 9 | 15.52 |

EC/TZ= end cervical/transformation zone component (EC/TZ); NILM= Negative for intraepithelial lesion or malignancy; ASCUS= Atypical squamous cells of undetermined significance; LSIL= Low-grade squamous intraepithelial lesion; HSIL= High-grade squamous intraepithelial lesion

**Figure 2:**
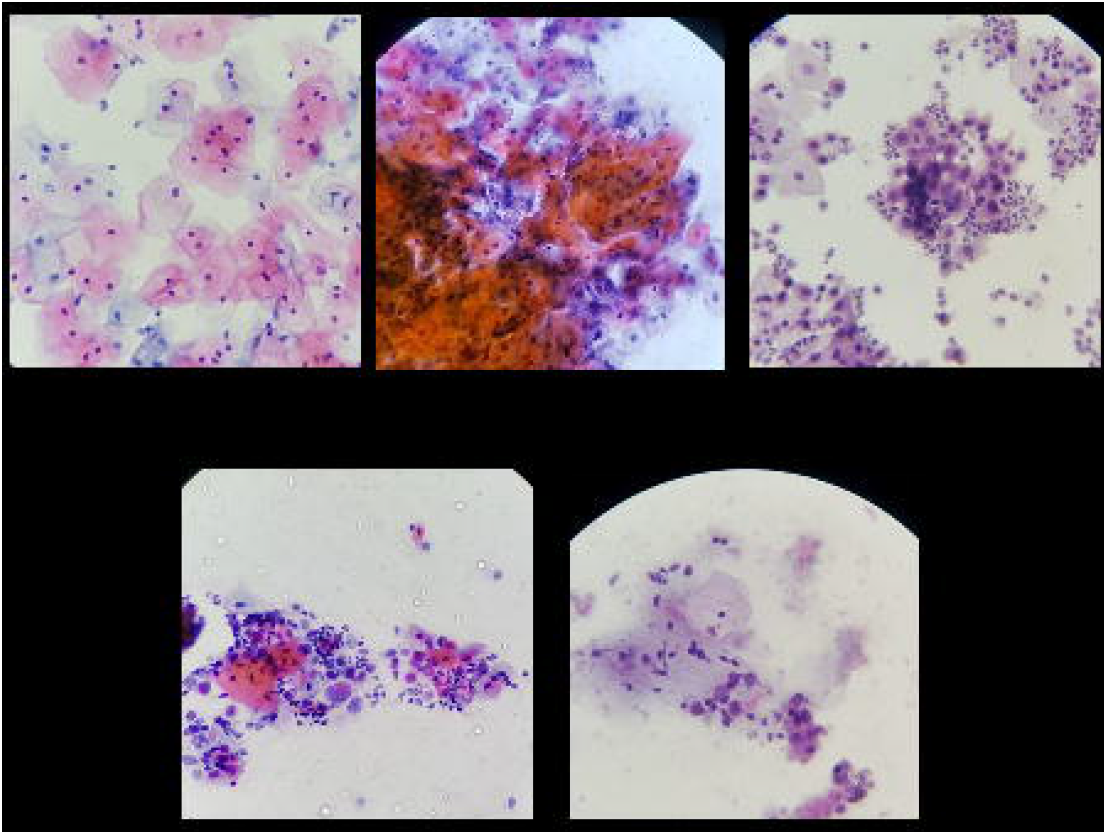
Papanicolaou smear showing a. Normal, b. Yeast-like Fungi, c. low-grade SILs, d. high-grade SILs and e. atypical squamous cells of undetermined significance lesions

#### Immunological status

CD4 counts was available for 57 patients. The mean CD4 count in the subjects was 593.66 ± 356.39 cells /μL (ranging from 20 to 1796). Approximately 60% had CD4 cell count above 500 cells/µL. We did not find a significant relationship between duration of disease, being under ART, marital status (married vs. unmarried), using the oral contraceptive pill, tubal ligation, and condom, with CD4 cells counts but age was negatively and statistically significant predictor of CD4 (*r*=0.33, P=0.01). However, the cytological abnormality was not related to the immunological status of patients indicated by CD4 cells count (see table 3).

**Table 3:** Association between CD4 count and presences of squamous cells abnormality (ASCUS and SIL), NILM and any abnormality.

| | $\beta$ | SEβ | P | Odd ratio (CI) |
| --- | --- | --- | --- | --- |
| ASCUS/SIL*, n= 17 | 0.009 | 0.038 | 0.81 | 1.01 (0.94-1.09) |
| NILM*, n= 41 | 0.063 | 0.039 | 0.11 | 1.06 (0.98-1.15) |
| Abnormal= 43 | 0.050 | 0.039 | 0.20 | 1.052 (0.97- 1.14) |

\*all models adjusted for age
ASCUS/SIL= atypical squamous cells of undetermined significance or squamous intraepithelial lesions; NILM= negative for intraepithelial lesions and malignancy; abnormal= presence of any cytological anomalies, organism, and inflammation

## 1. Discussion

In the current study, the prevalence of abnormal Pap smear in women with HIV was 29.3%, which is almost 6 time higher than in the general population. Comparable outbreak has been reported previously showing 26% cytological abnormal, but unlike our finding which ASC-US were the most common form of abnormality, they reported more LSIL lesions form[24]. In a study in Nigeria based on the Bethesda classification system, 27.6% of young HIV seropositive women had cervical abnormalities, which predominantly were LSIL (12.2%),[25]. Similar studies also reported that ECA is prevelant among HIV seropositive women ranging from 20 to 26%,[26, 27].

Various national and even local scale survey showed that prevalence of cervical abnormalities in HIV seropositive women is higher than normal population[28-37]. Therefore, it can be said that HIV is one of the independent risk factors for abnormalities and cervical cancer.

Our observation was n line with Gentient and Kodey studies which have demonstrated the most influenced sub-population were women above 30 years we have found that 70% of cervical abnormality apear to be among women aged 30 to 40 years[28, 30].

Also, it appears that 65% of smears were inflammatory and coexistences of inflammation and ASC-US observed in 81% of cases which is consistent with the results of other studies[28, 29]. Perhaps due to the smale number of observation alike to other studies we did not detected cervical cancer[29]. However many other studies showed an incidence of approximately equal to 0.1% during follow up period[30, 33, 34]

We did not find a link between the prevalence of Pap test abnormality and duration of disease or being under ART[27, 28] however, inconsistency exists in the literature[34].

We were unable to detect a statistically significant link between cervical squamous cell abnormalities and serum CD4 count. These results coincided with the findings of the study by Kodey et. all [28]. However, many studies have shown that HIV infected women with CD4 lower than 200 cells/μL or less than 350 are at high risk for cervical cell abnormalities[27, 30, 33, 34, 38-40]. This could be due to the fact that most of our patients had a CD4 count equal or more than 500 cells/µL and this probably lower power of our data to observe a risk difference if one existed.

in the current study although methodologically we obeyed from standard and globally reliable protocols and also controlled for many important individuals and environmental confounding factor, however, encountered some restrictions. A first limitation is a small number of samples thus; results should be interpreted with caution. In addition, because the study was conducted only in one center, the results of the present study cannot be generalized to the whole society.

High prevalence of abnormal Pap test among HIV seropositive women is a public health concern. Providing preventive measure such early detection of HPV as additional screening options in HIV-infected women is very important. Also needs for an updated guideline for cervical cancer screening and clinical management of the susceptible group.

## Data Availability

no additional data available

## Author Contributions

A.D. and S.S. researched, wrote, discussed and edited the manuscript. M.A.K analyzed, wrote and edited the manuscript. S.M.A. and A.S. contributed to the discussion and edited the manuscript. S.M. contributed to data collection. S.S. is the guarantor of this work and, as such, had full access to all the data in the study and takes responsibility for the integrity of the data and the accuracy of the data analysis.

## Data sharing

no additional data

## Ethical approval

the study was approved by the ethical committee, Deputy of Research and Development, Ahvaz Jundishapur University of Medical Sciences (IR.AJUMS.REC.1396.192).

## ACKNOWLEDGMENT

We acknowledge the contribution made by Mr. Homayon Amiri, Communicable Disease Prevention Unit, in data collection and data management.

The study was <u>supported by a grant</u> (Project no. OG-96105; Behsan ID 96362) from Research and Technology Deputy, Ahvaz Jundishapur University of Medical Sciences.

